# Improvement in quality of life and cognitive function in Post Covid Syndrome after online occupational therapy: results from a randomized controlled pilot study

**DOI:** 10.1101/2024.05.09.24307158

**Authors:** Dominik Schröder, Andrea Stölting, Christina Müllenmeister, Georg M.N. Behrens, Sandra Klawitter, Frank Klawonn, Aisha Cook, Nadja Wegner, Martin Wetzke, Tim Schmachtenberg, Alexandra Dopfer-Jablonka, Frank Müller, Christine Happle

**Author notes:** contributed equally (Dominik Schröder, Andrea Stölting, Alexandra Dopfer-Jablonka, Frank Müller, Christine Happle).

## Abstract

**Background:** Post-COVID-Syndrome (PCS) poses enormous clinical challenges. Occupational therapy (OT) is recommended in PCS, but structural validation of this concept is pending.

**Methods:** In an unblinded randomized pilot study (clinical trial # DRKS0026007), feasibility and effects of online OT in PCS were tested. Probands received structured online OT over 12 weeks either via interactive online treatment sessions (interactive group) or prerecorded videos (video group). 50% of probands received no online OT (control group). At week 0, 12, and 24, we analyzed study experience, health-related quality of life, and impairment in performance, participation, and cognitive functions.

**Results:** N=158 probands (mean age 38 yrs., 86% female) were included into the analyses. 83.3% of probands in the interactive versus 48.1% of probands in the video group described their study experience as positive or very positive (p=0.001). After 12 weeks, all groups displayed significant improvement in concentration, memory, and performance of daily tasks. After 24 weeks, significant improvement in concentration and memory were observed in control- and video-probands, and social participation had improved after video-OT. However, only probands that had received interactive online OT showed improvement of all measured endpoints including concentration, memory, quality of life, and social participation.

**Conclusion:** We show that online OT is feasible and that interactive online OT is a promising treatment strategy for affected patients. We present exploratory data on its efficacy and describe variables that can be employed for further investigations in confirmatory trials.

## Introduction

Approximately 3-10% of people with an acute SARS-COV-2 infection develop Post COVID-syndrome (PCS) with ongoing symptoms [1-3]. PCS affected people often experience severe fatigue, trouble concentrating, and reduced quality of life and social participation [4-6]. Current therapy guidelines for PCS mention occupational therapy (OT) as a treatment option [6], and case reports describe success of OT in treating this novel condition [7]. However, clinical studies confirming efficacy of OT in PCS are pending.

We sought to combine the concept of OT for PCS with the strategy of remote (digital) treatment delivery. Remote treatment strategies via digital communication have become more accepted since the COVID-19 pandemic [8]. As many PCS patients suffer from impaired mobility, digital treatment appears to be particularly appealing for this patient group [9]. Given the high prevalence of PCS, the scalability of digital therapy formats could enable efficient treatment for a high number of affected people.

Here, we tested the feasibility and efficacy of structured online OT for PCS in a randomized controlled, unpowered pilot study (German clinical trial registry #DRKS00026007 [10]). OT was based on a detailed manual tailored to the needs of PCS patients and delivered online, either by interactive sessions or as prerecorded videos. The aim of the intervention was to support affected people in developing coping strategies against major PCS symptoms such as fatigue and cognitive impairments and thus improve their quality of life and social participation.

## Methods

### Trial Design

This is a randomized controlled pilot study with two interventional groups receiving OT either via (I) interactive digital sessions (interactive group) or (II) prerecorded videos (video group). A control group (III) received no intervention (controls). Planned allocation ratio was 1:1:2 (I:II:III). Patients were evaluated upon study start (T1), after the 12-week treatment phase (T2) and 24 weeks after study start (T3). More study details are outlined in a study protocol [10]. No changes were made after trial commencement.

### Participants and Recruitment

Following inclusion criteria were applied: [1] age ≥ 16 years, [2] persistent or new PCS symptoms ≥ 4 weeks after SARS-CoV-2 infection (confirmed by PCR or rapid antigen testing), [3] feeling of strong cognitive impairment and/or fatigue (concentration deficits and/or fatigue) ≥ 5/10 on a Likert scale, [4] access to a digital device, [5] consent to participate in the study. Recruitment was performed through an online study platform for persons affected by PCS (DEFEAT Corona, German study registry number DRKS00026007 [11]). Interested patients were asked to complete an online screening survey. PCS people meeting the inclusion criteria were invited to participate by email. Prior to enrollment, individual consent talks were conducted which each interested eligible person and further study information was provided. Participants declared written consent before enrollment.

### Interventions

Participants in the interactive group received online OT by an experienced occupational therapist twice weekly through interactive digital sessions. Probands in the video group were provided with links for prerecorded OT videos for two weekly sessions. Outline and structure of OT were based on a detailed manual tailored to the needs of PCS patients which was identical in both interventional groups [10]. OT sessions consisted of guided exercises to control typical PCS symptoms as well as customizable units to improve performance in everyday occupations, social participation, and wellbeing. Participants in both groups also received OT workbooks and were asked to apply contents into their everyday life complementing the OT content of each therapy session regularly. Control participants received no treatment.

### Outcomes

Feasibility and acceptance of the treatment concept (primary endpoints) were analyzed by Likert scaled items (online questionnaire) and assessment of drop out rates in the different study phases. Secondary endpoints were cognitive function and problems in everyday occupations assessed by Neuro-QoL™ v2.0 cognitive function short form [12], memory function tested by the WIT-2 tool [13], and concentration deficits evaluated by the D2-R test [14]. Health-related quality of life was analyzed by the EQ-5D-3L index and EQ VAS [15]. Social participation was evaluated by the Index for measuring participation restriction (IMET) score [16], and occupational performance by the Canadian occupational performance measure COPM based interviews [13].

### Sample size

As pilot study, the sample size was conceived pragmatically considering feasibility with regard to project funding

### Randomization

Probands were randomized into the three groups employing an urn model (ratio interactive/video/control group: 1/1/2) by a member of the study team not involved in the treatment intervention. After randomization, patients were tested for occupational problems using a structured interview according to the Canadian occupational performance measure (COPM [13]), again by an independent team member not involved in the intervention.

### Statistical analyses

Sample characteristics where characterized using descriptive and bivariate statistics. Within groups, changes between two time points were assessed by Wilcoxon signed rank testing. To analyze longitudinal changes between groups, covariance analyses with repeated measurements were performed with baseline values, interventional arm, and time points as covariables. Analyses of treatment effects were conducted according to the intention to treat (ITT) principle [17], and missing values were imputed applying the last observation carried forward (LOCF) strategy [18]. Adjusted differences between interventional versus control groups at follow up visits are reported with confidence intervals (CI). P values were corrected for multiple testing by Bonferroni Holm correction, and a p value p<0.05 was considered statistically significant. Calculations were performed employing R (V4.2.3) with the packages afex, eq5d, and ggplot2 [10, 19, 20].

### Research Ethics

The study protocol was approved by responsible research ethics boards of all participating centers (Hannover Medical School #9948_BO_K_2021, University Medical Center Göttingen 15/8/22Ü). The study was registered at the German Registry for Clinical Trials (trial number DRKS00026007). Participants provided written informed consent prior to enrollment.

The project received funding through the German Federal Ministry for Education and Research (Grant number 01EP2103C).

## Results

Initially, n=163 PCS patients were recruited, but n=5 had to be excluded secondarily due to failing the inclusion criteria (Fig. 1). In total, n=158 PCS patients were included into the final analysis. 65.2% of participants completed the 12-week interventional or control period and were available for evaluation and timepoint T2. 32.5% of probands in the interactive, and 53.8% of participants in the video group were lost to follow up at T2, as were 22.8% of control probands. An additional 2.5% of participants were lost to follow up at timepoint T3.

**Fig. 1:**
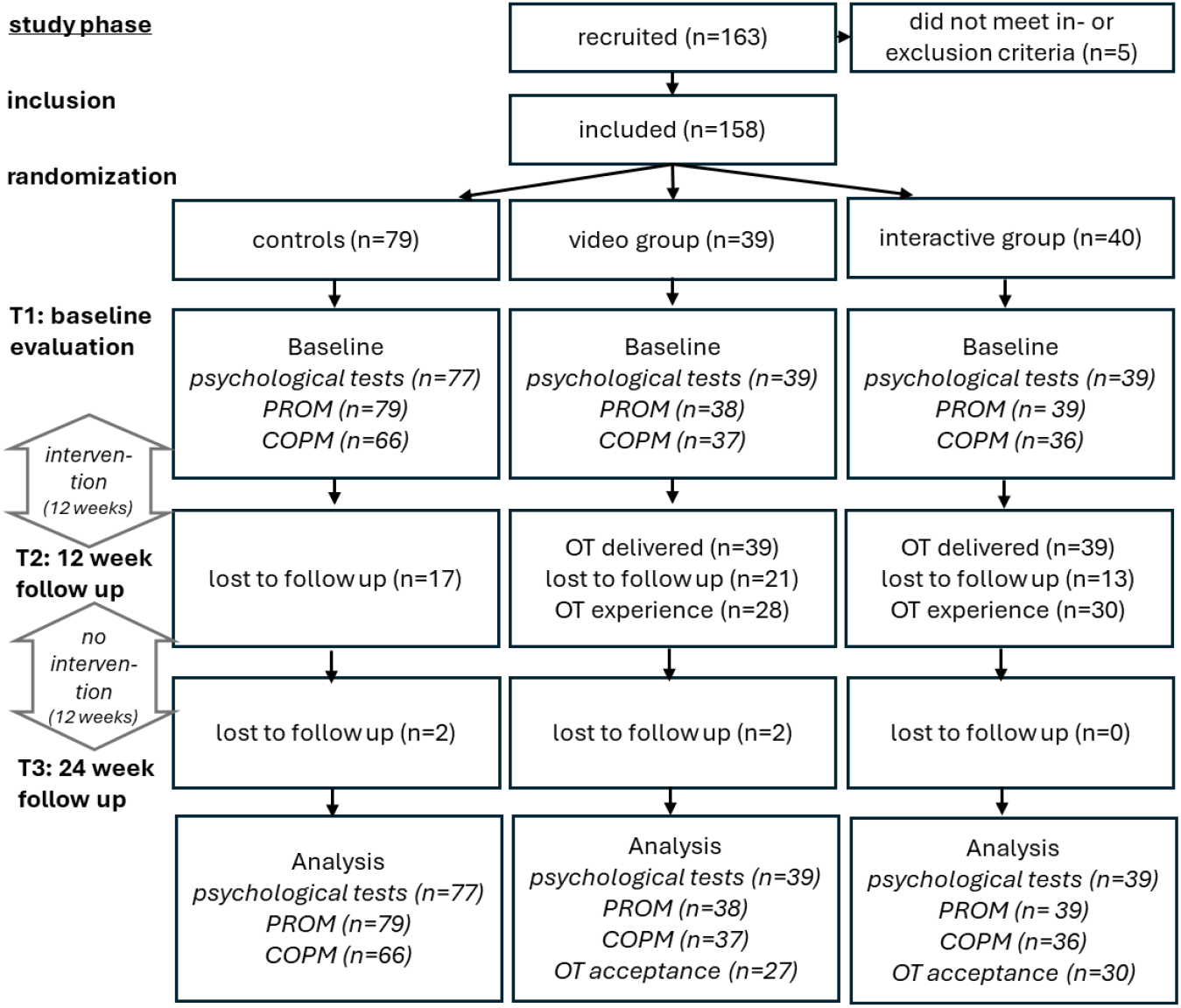
Flowchart illustrating study phases and proband numbers (PROM: patient reported outcome measures include NeuroQoL, EQ-5D, IMET)

From cases in the interventional arms with follow up data, data on subjective treatment effects was available for 98.7% of probands, and 72.2% provided information on their personal OT experience. Psychological testing was performed in 98.1%, and COPM in 88% of probands with follow up evaluation. Participant numbers for each study arm and timepoint are illustrated in Fig. 1.

The median age of participants was 38 years (IQR 30–45, range 16–67), and probands in the interventional groups were significantly younger than those in the control group (Table 1). The majority of participants (86.1%) were female, with more women in the interventional as in control groups. PCS participants in the interactive group presented with better memory performance (WIT-2) than those in control and video arms. Other characteristics that did not differ between the study groups are displayed in Table 1. Further information on sociodemographic characteristics is provided in supplementary Table 1.

**Table 1:** Proband characteristics at baseline.

|  | Total<br>(n=158) | Control<br>(n=79) | Video<br>(n=39) | Interactive<br>(n=40) | p-value |
| --- | --- | --- | --- | --- | --- |
| Age<br>(years) <sup>a</sup> | 38.0 (30.0 – 45.0)<br>*Range 16 – 67* | 42.0 (31.5 – 47.0)<br>*Range 16 – 61* | 35.0 (26.5 – 41.0)<br>*Range 18 – 53* | 36.0 (29.0 – 44.0)<br>*Range 16 – 67* | 0.038<sup>1</sup> |
| Female gender | 136 (86.1) | 51 (82.3) | 24 (88.9) | 17 (94.4) | 0.025<sup>2</sup> |
| SARS-CoV-2 infection<br>until study start (weeks,<br>n=5 missing) | 41.1 (26.8 – 63.9) | 43.1 (28.1 – 60.9) | 38.0.7 (20.9 – 51.2) | 42.6 (28.9 – 98.3) | 0.251<sup>1</sup> |
| Fatigue<sup>a</sup><br>(Likert x/10) | 8.0 (7.0 – 9.0) | 8.0 (7.0 – 9.0) | 9.0 (7.0 – 10.0) | 8.0 (7.0 – 9.0) | 0.650<sup>1</sup> |
| Trouble concentrating<br>(Likert x/10)<sup>a</sup> | 8.0 (7.0 – 9.0) | 8.0 (7.0 – 9.0) | 8.0 (7.0 – 9.0) | 7.0 (6.0 – 9.0) | 0.757<sup>1</sup> |
| d2-R<sup>a</sup><br>(n=3 missing) | -1.0 (-1.6 – -0.2) | -1.1 (-1.0 – -0.2) | -0.9 (-1.6 – -0.2) | -0.8 (-1.5 – -0.1) | 0.394<sup>1</sup> |
| WIT-2<sup>a</sup><br>(n=3 missing) | 0.1 (-0.6 – 0.5) | -0.1 (-0.5 – 0.3) | -0.1 (-1.0 – 0.3) | 0.5 (-0.2 – 0.9) | 0.007<sup>1</sup> |
| EQ-5D-3L Index<sup>a</sup> (n=2<br>missing) | 0.61 (0.39 – 0.75) | 0.61 (0.39 – 0.75) | 0.61 (0.38 – 0.75) | 0.61 (0.484 – 0.75) | 0.640<sup>1</sup> |
| EQ VAS<sup>a</sup><br>(n=2 missing) | 36.0 (30.0 – 51.0) | 37.0 (30.0 – 50.0) | 35.5 (30.3 – 49.0) | 36.0 (31.0 – 56.0) | 0.576<sup>1</sup> |
| Neuro-QoL™ v2.0<br>Cognitive Function-Short<br>Form<sup>a</sup> (n=3 missing) | 33.0 (29.8 – 36.0) | 34.0 (29.8 – 36.0) | 33.0 (28.6 – 37.0) | 32.0 (28.6 – 35.0) | 0.833<sup>1</sup> |
| IMET<sup>a</sup><br>(n=6 missing) | 55.0 (41.3 – 66.0) | 53.5 (41.0 – 66.0) | 54.5 (44.8 – 66.0) | 59.5 (37.8 – 66.0) | 0.893<sup>1</sup> |
| COPM – satisfaction<br>(n=19 missing) | 2.3 (1.5 – 3.0) | 2.3 (1.5 – 3.0) | 2.5 (1.3 – 3.0) | 2.4 (1.5 – 3.3) | 0.636<sup>1</sup> |
| COPM – performance<br>(n=19 missing) | 3.0 (2.5 – 3.6) | 3.0 (2.5 – 3.5) | 3.0 (2.5 – 3.8) | 3.0 (2.5 – 3.8) | 0.816<sup>a</sup> |

<sup>1</sup>Kruskal-Wallis testing; <sup>2</sup>Fisher-Freeman-Halton testing; <sup>a</sup>median (IQR); <sup>b</sup>number (proportion); <sup>\*</sup>comparison between control, video, and interactive group

To further analyze the acceptance of our concept, we asked PCS patients to rate their OT experience during the 12-week intervention phase. In an online survey, they could rate their therapy perception between 0 (very negative) and 5 (very positive). Participants in the interactive group evaluated their OT experience with a mean of 4.2 points (range 2–5, n=30), and 83.3% rated the treatment as positive or very positive (Fig. 2). By contrast, probands in the video group rated their OT experience with an average of 3.3/5 points (range 1–5, n=27), and only 48.1% described their experience as positive or very positive (Fig. 2, difference in satisfaction p=0.001).

**Fig. 2:**
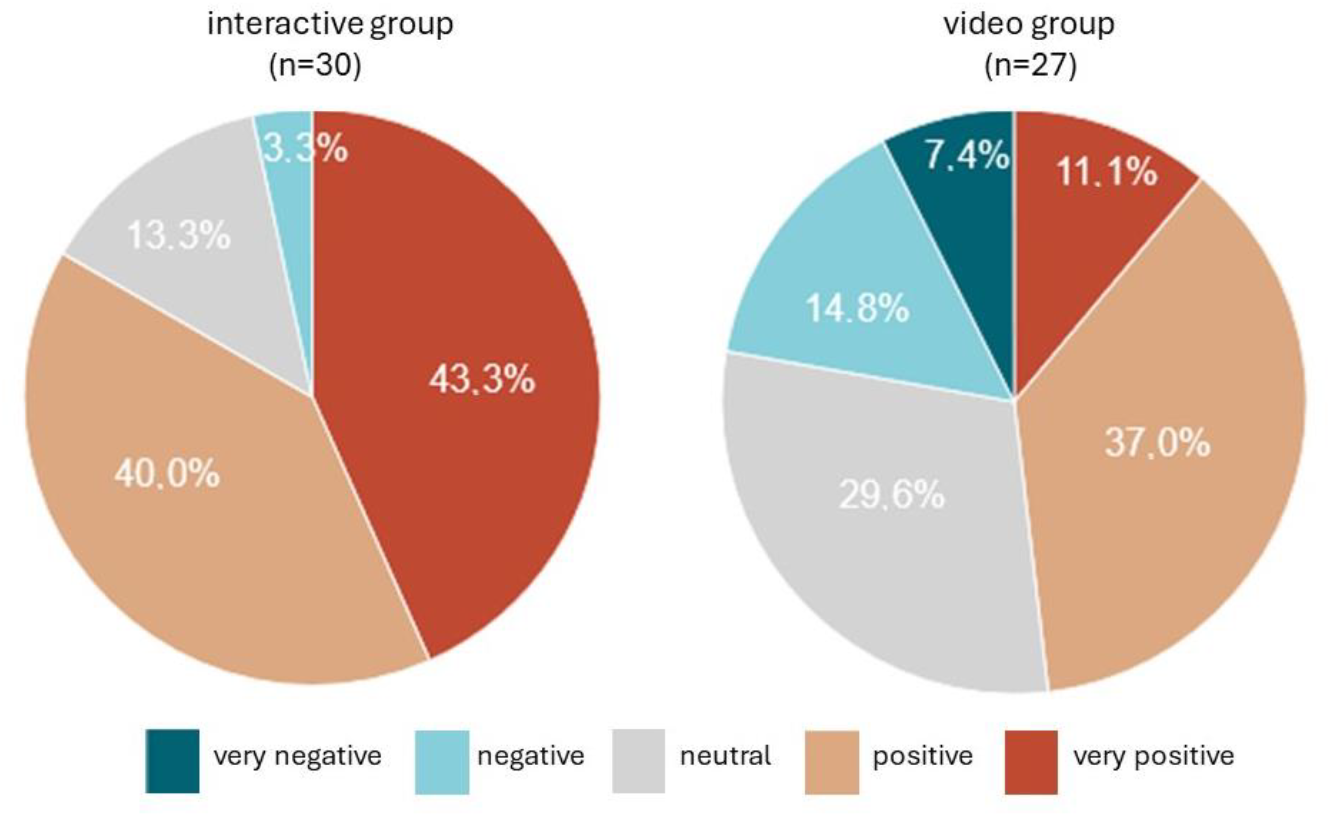
Rating of study experience by participants in the two intervention groups.

Next, we performed an exploratory analysis of the intervention’s efficacy in alleviating PCS symptoms. In all groups, a significant improvement of concentration and memory, as well as occupational performance was observed during the first 12 weeks of the study (Table 2). However, only in participants after interactive online OT, a significant improvement in health-related quality of life, social participation, and cognitive function as assessed by Neuro-QoL occurred (Table 2). In comparison to controls, participants in the interactive group presented with significantly stronger improvement in health-related quality of life (EQ-5D-3L index, mean difference 0.11, 95% CI [0.02-0.19], p=0.01) and cognitive function (Neuro-QoL, mean difference 2.36, 95% CI [0.27-4.46], p=0.022, Table 2).

**Table 2:**
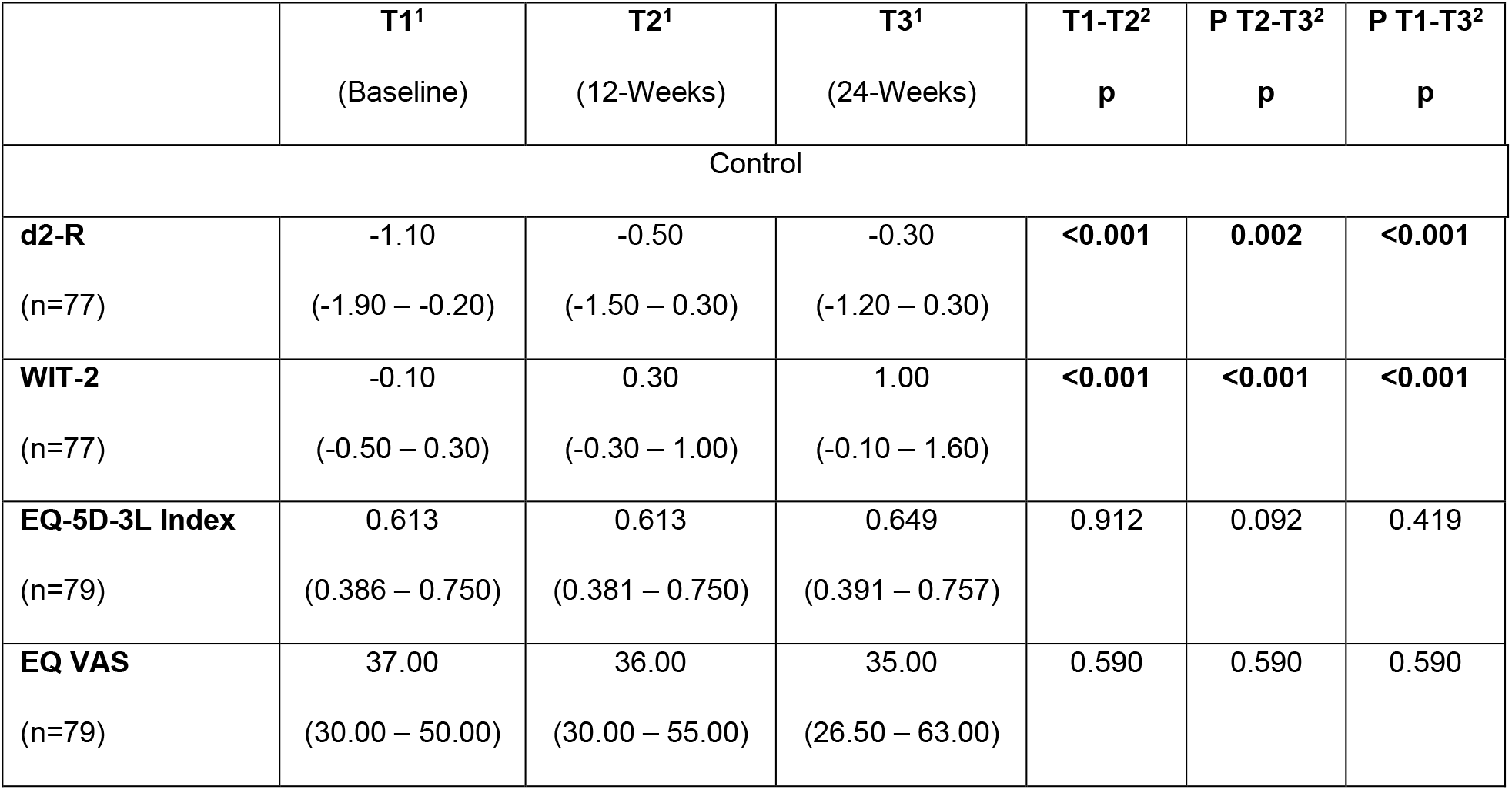
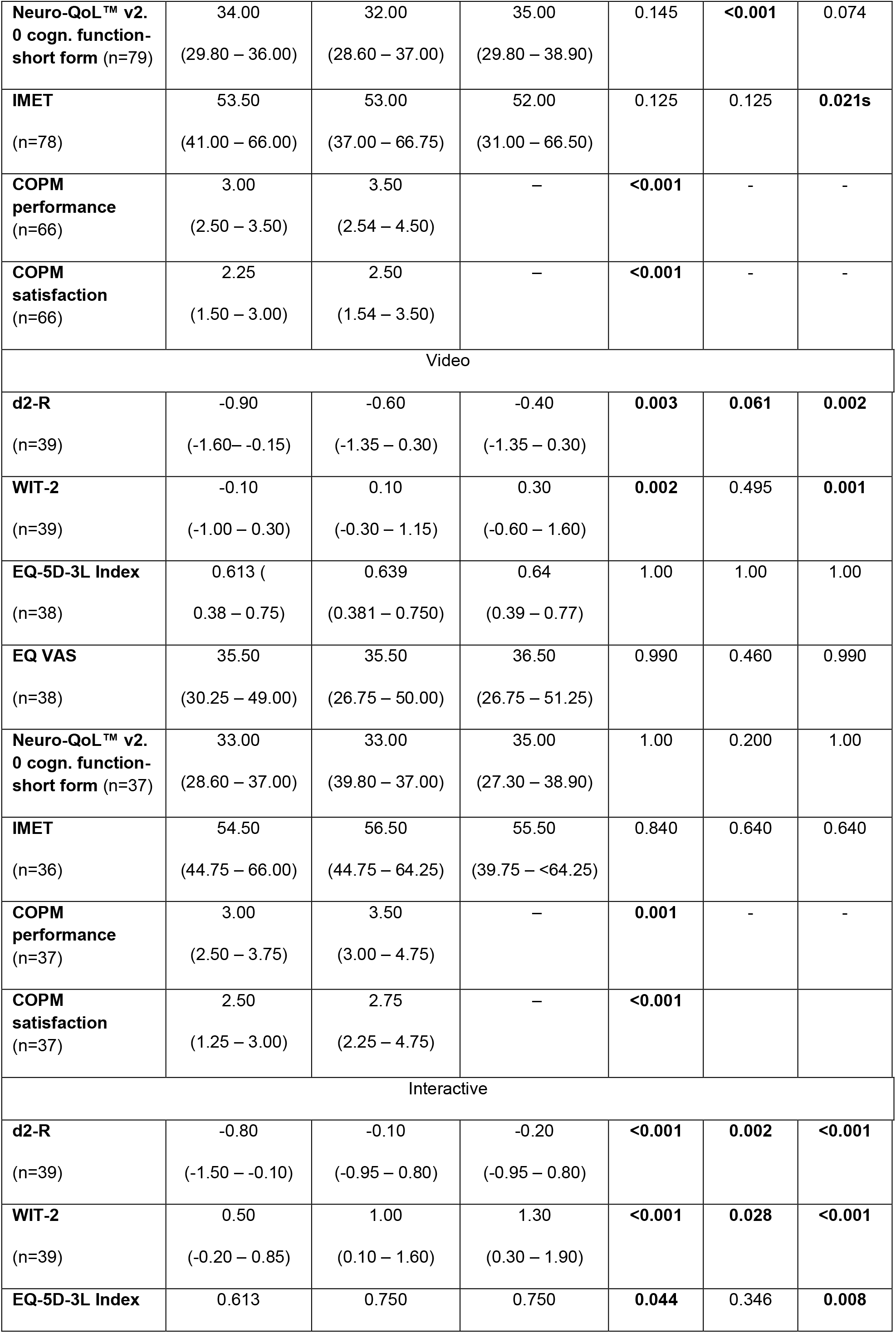
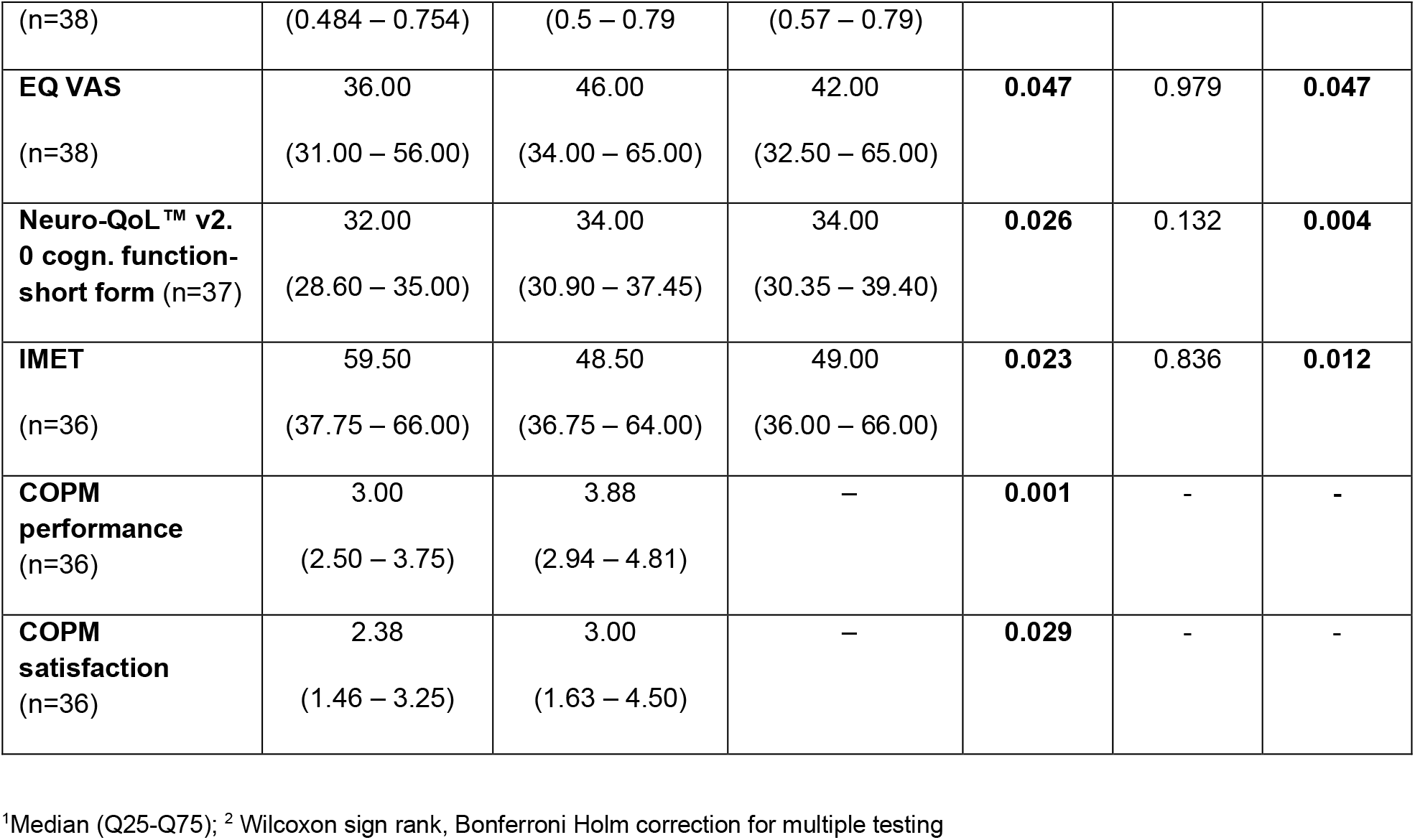
Effects of the study intervention on cognitive performance, quality of life, and occupational performance.

24 weeks after the start of the study, all groups displayed significant improvement in concentration and memory performance, and participants in the control and interactive groups showed significant enhancement of social participation (Table 2). However, only PCS patients in the interactive group presented with significant improvement of all measured end points such as cognitive function, social participation, health-related quality of life, and occupational performance (Table 2).

## Discussion

PCS is a growing global health problem which has been estimated to affect up to 11% of patients after SARS-CoV-2 infection [3]. In spite of high patient numbers worldwide and strong clinical need, effective treatment strategies are pending [21, 22].

Here, we present first results on the feasibility of online OT and explore the efficacy in PCS. The delivered parameters and data can be employed in larger randomized controlled clinical trials on novel PCS treatment strategies. Our data illustrates that online OT can be a valuable treatment approach in helping PCS patients better coping with their symptoms and restraints. We have shown that online delivered OT can significantly improve their cognitive function and quality of life.

Online delivered OT has several advantages especially for people with PCS: It can easily be scaled meeting the current demand for treatment and may also be especially suitable for PCS people that struggle with impaired mobility or live in more remote places. In our study, online provision of OT was used by participants of all ages, illustrating the wide range of PCS patients that could be reached by such an approach. Around two thirds of PCS patients in the interactive therapy group completed the 12-week treatment course in the interactive group as compared to less than one half of those in the video group, and significantly more patients that were treated interactively rated their OT experience as positive. This illustrates that personal interaction significantly enhances acceptance of online treatment in PCS. Furthermore, PCS patients after interactive OT displayed significant treatment benefits when compared to both video and control group. Their cognitive function showed statistically significantly improvement and their health-related quality of life had increased by 0.11 points, a value clearly above the threshold of 0.03 previously described to be clinically relevant [23]. The fact that secondary outcomes such as memory and concentration capacity also improved in the control group could be explained by a natural amelioration of PCS symptoms over time, which has also been observed by other researchers [24]. Importantly, only patients receiving interactive OT showed significant improvement of all measured endpoints of cognitive function, occupational problems, quality of life, and social participation.

Taken together, these findings advocate for high acceptance and therapeutic benefit of the interactive, person to person online OT in PCS patients.

Our study has several limitations. Firstly, due to it’s novelty, *a priori* power calculations could not be performed and the number of enrolled participants may have been too low to detect subtle treatment effects. Also, the inhomogeneity of groups upon study start with regard to age, sex, and memory function may have impacted our results. The high rate of lost to follow up participants needs to be addressed in future studies. Unfortunately, we were unable to systematically assess reasons for study discontinuation in our cohort. The high proportion of female study participants can be interpreted as another limitation of our work. It may be explained by a higher rate of female PCS patients in general and the fact that women tend to seek medical help earlier than men [25, 26]. Furthermore, central outcomes such as symptom severity were self-reported. As this feasibility study was performed in unblinded fashion, this could have biased self-assessment. Finally, some of our test instruments such as WIT-2 and d2r are not validated for repeated measurements, which could have led to improvement of scorings at the second test date.

In spite of these limitations, our study adds significant information to the field of PCS treatment. We provide detailed data on relevant clinical variables that can be employed in larger studies on PCS treatment in the future. Our exploratory analyses of treatment effects also support the notion that online OT is feasible in PCS and that interactive online OT may be a promising treatment strategy for affected patients.

As such we hope our data helps to pave the way for new, effective, and scalable treatment options in PCS.

## Data Availability

Data cannot be shared publicly because they contain personal information. Data are available from the Hannover Medical School Institutional Data Access (contact via Study director PD Dr. med. Alexandra Jablonka Clinic for Immunology and Rheumatology - OE 6830 Hanover Medical School Carl Neuberg Strasse 1 30625 Hannover Phone: +49 511 532 3014) for researchers who meet the criteria for access to confidential data.

## Supplement

**Supplementary Table 1:** Differences between study arms adjusted for baseline scores.

|  | T2 difference<br>(95% CI)<sup>1</sup> | T3 difference<br>(95% CI)<sup>1</sup> | P T2<sup>2</sup> | P T3<sup>2</sup> |
| --- | --- | --- | --- | --- |
| d2-R<sup>3</sup> |  |  |  |  |
| video vs. control | -0.07 (-0.42 – 0.28) | -0.14 (-0.57 – 0.30) | 1.00 | 1.00 |
| interactive vs. control | 0.08 (-0.28 – 0.43) | 0.15 (-0.28 – 0.59) | 0.783 | 0.783 |
| WIT-2<sup>3</sup> |  |  |  |  |
| interactive vs. control | 0.07 (-0.31 – 0.46) | -0.41 (-0.92 – 0.10) | 1.00 | 1.00 |
| video vs. control | 0.10 (-0.28 – 0.49) | -0.18 (-0.70 – 0.32) | 0.158 | 0.746 |
| EQ-5D-3L index<sup>3</sup> |  |  |  |  |
| video vs. control | 0.021 (-0.069 – 0.110) | 0.006 (-0.108 – 0.095) | 0.577 | 0.880 |
| interactive vs. control | 0.106 (0.020 – 0.193) | 0.065 (-0.033 – 0.163) | **0.011** | 0.332 |
| EQ VAS<sup>3</sup> |  |  |  |  |
| video vs. control | -1.21 (-8.47 – 6.05) | -4.47 (-13.27 – 4.32) | 0.685 | 0.655 |
| interactive vs. control | 4.09 (-3.89 – 12.07) | 0.32 (-9.35 – 9.98) | 0.449 | 0.936 |
| Neuro-QoL™ v2.0 cognitive function short form<sup>3</sup> |  |  |  |  |
| video vs. control | 0.48 (-1.63 – 2.58) | -1.25 (-4.26 – 1.77) | 0.582 | 0.633 |
| interactive vs. control | 2.36 (0.27 – 4.46) | 0.80 (-2.20 – 3.81) | **0.022** | 0.633 |
| IMET<sup>4</sup> |  |  |  |  |
| video vs. control | 3.78 (-4.17 – 11.74) | 3.63 (-5.67 – 12.93) | 0.498 | 0.686 |
| interactive vs. control | -3.40 (-11.54 – 4.74) | -2.31 (-11.82 – 7.21) | 0.498 | 0.686 |
| COPM performance |  |  |  |  |
| video vs. control | 0.13 (-0.50 – 0.75) | - | 1.00 | not assessed |
| interactive vs. control | 0.20 (-0.46 – 0.85) | - | 1.00 | not assessed |
| COPM satisfaction |  |  |  |  |
| video vs. control | 0.30 (-0.45 – 1.05) | - | 0.977 | not assessed |
| interactive vs. control | 0.10 (-0.74 – 0.94) | - | 1.00 | not assessed |

<sup>1</sup>differences and CI adjusted according to baseline values employing repeated measurement variance analyses, <sup>2</sup>Bonferroni Holm adjusted for multiple testing, <sup>3</sup>positive value: improvement, <sup>4</sup> negative value: improvement.

